# Factors affecting anal sphincter recruitment during intraoperative pudendal nerve stimulation: an observational study

**DOI:** 10.1101/2024.06.14.24308947

**Authors:** Amador C Lagunas, Po-Ju Chen, Luis Ruiz, Amolak S Jhand, Nystha Baishya, Scott Lempka, Priyanka Gupta, Tim Bruns

**Author notes:** **Corresponding author:** Tim M. Bruns, Address: B10-A169 NCRC, 2800 Plymouth, Ann Arbor, MI 48109-2800.

## Abstract

**Introduction and Hypothesis:** The relationship between pudendal neuromodulation and patient factors is not well understood. This observational study aimed to identify and quantify physiological, demographic, and stimulation factors that significantly affect external anal sphincter (EAS) recruitment and outcomes in participants receiving pudendal neuromodulation for treatment of lower urinary tract symptoms and pelvic pain.

**Methods:** Participants (N=16) provided demographic and diagnostic information upon entry to this observational study. EAS activation at different stimulation amplitudes and pulse widths was recorded during lead implantation. Magnetic resonance imaging and computed tomography were used to determine the distance of the electrodes on the implanted lead from the nerve. Linear mixed modeling was used to quantify the impact of each variable on EAS recruitment.

**Results:** Participant sex, age, and body-mass index did not significantly affect EAS recruitment. Participant diagnoses had significant relationships to EAS recruitment, likely due to unbalanced group sizes. A pulse width of 210 µs required less current than 60 µs (p = 0.005) and less charge than 450 µs (p = 0.02) to activate the EAS. Increased electrode-to-nerve distance decreased the magnitude of the EAS response (p = 0.0011), increased the EAS activation threshold (p < 0.001), and was related to reduced bladder symptom improvements.

**Conclusions:** Of the three tested pulse widths, 210 µs best balances current and charge for EAS recruitment. Minimizing the distance between the electrode and pudendal nerve should be a priority during lead implantation. External sphincter activation threshold and response magnitude could be useful clinical indicators of electrode-to-nerve distance.

## Introduction

Chronic pelvic pain and lower urinary tract disorders (LUTD) affect millions of people in the United States [1,2]. These conditions result in diminished physical health, mental health, and productivity [3–5] placing a large and growing burden on the healthcare system [6]. Therapy options for chronic pelvic pain and LUTD are typically grouped on their level of invasiveness including non-invasive, pharmacologic, and minimally invasive therapies like botulinum toxin injections and sacral neuromodulation [7].

When patients fail to respond to traditional therapies they are left with few options. Pudendal nerve stimulation has emerged as a minimally invasive off-label therapy for pelvic pain and LUTD, successfully improving symptoms even in people who previously failed sacral neuromodulation[8,9]. While pudendal neuromodulation often improves symptoms, outcomes can vary and not all patients gain complete relief [10–13]. These differences in outcomes could be due to variations in patient anatomy, altered LUT function due to their symptom etiology, or treatment factors such as the distance between the electrode and nerve. Aging can change nerve function[14] and therefore effectiveness at tolerable stimulation amplitudes, while body composition may affect the electrode-to-nerve distance and thus nerve recruitment with electrical stimulation. Insights into the factors which have the largest effect on nerve activation and treatment outcomes may further improve pudendal neuromodulation outcomes.

Typically, the neurostimulator implant procedure involves two surgical stages. In the stage-1 implant surgery, the stimulation lead is placed near the main trunk of the pudendal nerve and the pulse generator is placed in a later stage-2 implant surgery. Anal sphincter recruitment in response to pudendal stimulation during the stage-1 surgery is a key indicator that the lead is correctly positioned and recruiting fibers in the main trunk of the pudendal nerve [15].

However, to our knowledge, no previous study has investigated what factors influence recruitment of the anal sphincter. Our primary objective was to determine which stimulation parameters and physiological factors have a significant impact on the recruitment of the inferior rectal branch of the pudendal nerve during pudendal nerve stimulation. Our primary hypotheses were that smaller electrode-to-nerve distances, longer stimulation pulse widths, and larger stimulation amplitudes would increase pudendal nerve recruitment, as observed via anal sphincter activity. We also hypothesized that smaller electrode-to-nerve distances and longer stimulation pulse widths would reduce nerve activation thresholds. Finally, we hypothesized that participant demographics and lower urinary tract symptoms would not be significantly related to nerve recruitment or thresholds.

## Materials and Methods

Approval for this observational study was obtained from the Institutional Review Board (HUM00165005). This study was registered on ClinicalTrials.gov (NCT04236596) before commencement of study procedures. This was a single center study with all surgeries performed by the same physician.

We recruited patients 18 years of age or older receiving pudendal neuromodulation for the treatment of bladder dysfunction and/or pelvic pain. Individuals were excluded from the study if they were pregnant or planned to be pregnant during the study or were diagnosed with atonic bladder, neurogenic bladder, pudendal nerve damage, or other conditions affecting the neural circuits in control of micturition. Individuals were also ineligible if they were unable to provide consent or unable to complete all study activities. Participants were recruited from September 2020 to June 2023. We targeted a sample size of 20 participants based on the expected number of pudendal implants in our clinical practice within that timeframe and similar human subject neuromodulation studies [16].

After providing informed consent, participants completed clinically validated surveys to assess bladder health (American Urological Association Symptom Index: AUASI [17] and Michigan Incontinence Symptom Index: M-ISI [18]), sexual function (Female Sexual Function Index-6: FSFI-6 [19] or Sexual Health Inventory for Men: SHIM [20]), bowel function (Colorectal-Anal Distress Inventory-8: CRAD-8 [21]), and pelvic pain (Female Genitourinary Pain Index: FGUPI [22] or Male Genitourinary Pain Index: MGUPI [22]). Just before or after the final post-op clinic visit participants completed the same set of surveys, and we calculated survey score changes. Participants underwent a pre-surgery pelvic magnetic resonance imaging (MRI) scan, data collection during their scheduled stage-1 implant surgery, and a pelvic computed tomography (CT) scan at least three weeks after the stage-2 surgery.

Data was collected during the stage-1 surgery, in which an Interstim (Medtronic, MN, USA) electrode lead was inserted near the pudendal nerve, as described in Peters 2013 [23], using fluoroscopy and external anal sphincter (EAS) electromyography (EMG) to confirm lead position. Monopolar needle EMG electrodes (Ambu, MD, USA), were placed in the external anal sphincter at the 3 and 9 o’clock positions. The needle electrodes were connected to a Cascade Elite (Cadwell, WA, USA) intraoperative neurophysiological monitor on which the EAS EMG data was recorded with a 333 hertz (Hz) sampling rate. The EMG data was filtered by the Cascade system with 3 kHz low pass and 10 Hz high pass filters, and a 60 Hz notch filter when 60 Hz noise was observable.

After lead placement, 10-second stimulation trials were performed in which the stimulation amplitude, pulse width (among 60, 210, and 450 µs), and electrode combinations were varied. The implanted lead had four electrodes (E0 to E3) that could be activated individually (monopolar referenced to a surface electrode on the ankle) or used in any bipolar combination once the external pulse generator was connected. Stimulation was performed at either 3 Hz for monopolar stimulation or 3.1 Hz for multipolar stimulation. For each electrode and pulse width combination, threshold amplitude was defined as the lowest stimulation amplitude that elicited an EAS contraction. Stimulation trials were performed at and above the threshold amplitude to a maximum of 5.5 mA. Charge per pulse was calculated by multiplying the applied current by the pulse width for a given trial.

We used volumetric imaging from both the pre-surgery MRI (3T T2 weighted pelvis scan; Ingenia 3T system, Philips, Amsterdam, the Netherlands) and post-surgery helical CT scans (L4 vertebra to the proximal femur shaft with a slice thickness of 0.625 mm; Discover CT750 HD or Revolution scanner, GE Healthcare, IL, USA) together to create patient-specific models of the pelvic anatomy. We used Mimics Research (v25.0; Materialise, Leuven, Belgium) and the Vascular Modeling Toolkit (v1.4.0; http://www.vmtk.org/) to identify and segment the pudendal nerve in the MRI scans and to identify and segment the implanted lead artifacts in the CT scan. We then used Mimics Research to co-register surface segmentations derived from the MRI and CT scans and to position the lead and nerve within the pelvis. We recreated the lead geometry from the CT artifacts using the geometry functions of Comsol Multiphysics (v 5.6; Comsol, Stockholm, Sweden). To measure the electrode-to-nerve distance we calculated the center-to-center distances between each electrode on the implanted lead and the closest point on the pudendal nerve, in Python (v3.7.0), using their recorded positions from the CT and MRI scans respectively. We did not blind participant study identifiers or sex during image analysis, however researchers performing segmentation were unaware of the intraoperative data collection outcomes for each participant.

For each stimulation trial, we calculated the average area under the curve (AUC) of the rectified EAS EMG for five stimulation events using MATLAB (v2021a, MathWorks, Natick, Massachusetts, USA). This value was used to quantify the magnitude of the anal sphincter contraction, as done previously [24–26]. The AUC calculation interval started 10 milliseconds after the stimulation artifact onset and continued for 100 milliseconds. This process is visually summarized in Supplemental Figure 1.

We performed all statistical analysis with R Statistical Software (v4.4.1; R Core Team 2024). We used a Shapiro-Wilk test to check for normality of each data set. A Kruskal-Wallis test was used to determine if groups had equal medians followed by a pairwise Mann-Whitney U test with a Bonferroni correction to determine statistically significant differences between groups. Where relevant, values are given for the median and interquartile range (IQR) or average and standard deviation. A significance level of α = 0.05 was used. We used linear mixed models to estimate the EAS AUC and the threshold current based on participant age, sex, body mass index (BMI), lower urinary tract symptoms (overactive bladder (OAB), urinary retention, urinary incontinence, and pelvic pain), and stimulation parameters (stimulation amplitude, pulse width, electrode-to-nerve distance). We only included participants who had a lead migration distance of 5 mm or less in models incorporating electrode-to-nerve distance data, based on a comparison of each participant’s stage-1 fluoroscopy images and CT images. We chose this distance because adjacent electrodes on the lead are roughly 5 mm apart. We used the R packages *lme4* and *lmerTest* to calculate the linear mixed model parameters. In the model, pulse width is a 3-level categorical variable with a 60 µs pulse width used as the baseline condition. Nerve-to-electrode distance, stimulation amplitude, age, and BMI were used as continuous variables with all other variables included as yes/no categoricals. ANOVA was used to compare models and determine which fixed effects to include. Participants were classified as having OAB, retention, incontinence, or pelvic pain based on the diagnoses given by their examining urologist. Diagnoses of urge urinary incontinence, stress urinary incontinence, or mixed urinary incontinence were all classified as incontinence. Diagnoses of pelvic pain, pudendal neuralgia, vulvodynia, and interstitial cystitis were all classified as pelvic pain in the statistical analysis.

## Results

Of the 23 participants eligible for study inclusion, seven did not complete intraoperative data collection for a variety of reasons including: improvement of symptoms eliminating the need for a neurostimulator, insurance denial or delay, unsuccessful device implantation, inability to record data due to COVID restrictions, indefinite surgical postponement, or non-signature of consent form and were not included in this analysis. We did not observe a relationship between these factors and patient demographics or collected data. Patient information for the included sixteen participants is summarized in Table 1. One female participant had their implanted lead removed and was reenrolled.

**Table 1.** Summary of participant characteristics. Average and (standard deviation) given when relevant.

| Variable | Count |
| --- | --- |
| Sex |  |
| Female | 13 |
| Male | 3 |
| Diagnosed Condition |  |
| Overactive Bladder | 12 |
| Retention | 5 |
| Incontinence | 7 |
| Pelvic Pain | 12 |
| Age (years) | 55.9 (10.8) |
| BMI (kg/m<sup>2</sup>) | 28.4 (3.8) |

BMI: body mass index

The monopolar EAS threshold current for 210 and 450 µs pulse widths were not normally distributed (p < 0.01) and had a median of 1.90 (IQR = 1.2), 0.75 (IQR = 0.45), and 0.55 (IQR = 0.45) mA for a pulse width of 60, 210, and 450 µs, respectively. The three pulse widths had significantly different (p = 0.001) median threshold levels, with the threshold current level at 60-µs significantly higher than 210 µs (p = 0.005) and 450 µs (p = 0.004) (Figure 1A). Similarly, the charge per pulse at EAS threshold was not normally distributed (p < 0.001). The median charge required for 60, 210, and 450 µs pulse widths was 114.0 (IQR = 72.0), 157.5 (IQR = 94.5), and 247.5 (IQR = 202.5) nC, respectively. These three groups did not have the same median (p = 0.001). There was a significant difference between the median charge required at anal sphincter threshold for 450 and 210 µs pulse widths (p = 0.02) and 450 and 60 µs pulse widths (p = 0.004) (Figure 1B).

**Figure 1.**
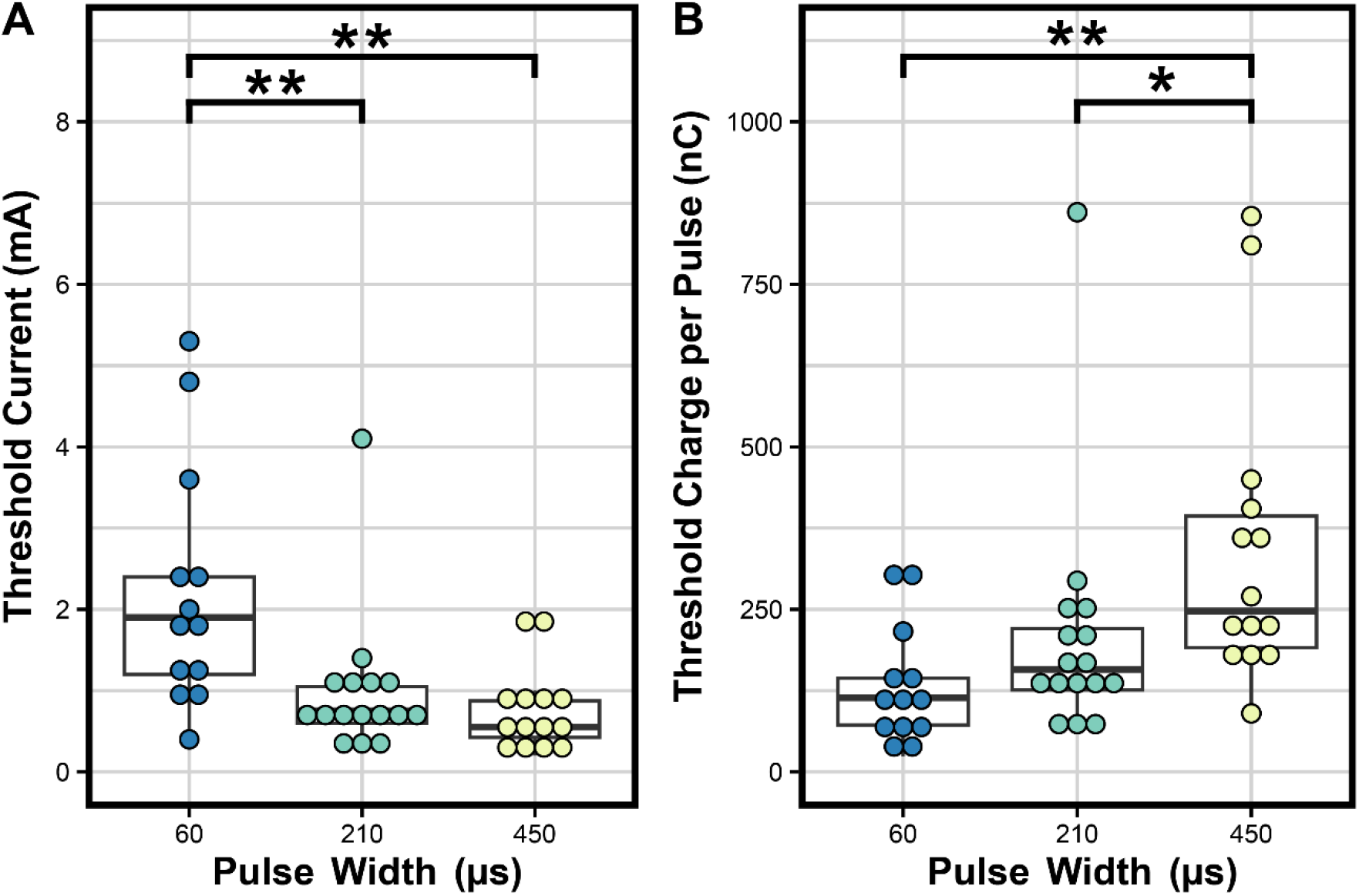
Characterization of external anal sphincter EMG activation by intraoperative monopolar pudendal nerve stimulation. Each dot represents the minimum threshold across all electrodes for an individual participant at a given pulse width. A) The threshold current required to activate the EAS for the tested pulse widths of 60, 210, and 450 µs. B) The charge per pulse at the EAS activating threshold current depending on pulse width. Box plots represent the IQR for the height of each box, with the bold central line indicating the median. Tips of whiskers extend to the value(s) within 1.5 x IQR of each box side. * indicates p < 0.05, ** indicates p < 0.01.

Supplemental Figure 2 provides individual participant monopolar threshold current and charge levels for each tested pulse width. In this figure it is shown that at a 210 µs pulsewidth 15 of 16 participants had EAS activation below 2 mA. There was not a significant difference between monopolar and bipolar thresholds (p = 0.98) (Supplemental Figure 3).

In a linear mixed model incorporating all monopolar trials, the estimated AUC was significantly impacted by stimulation current, stimulation pulse width, and diagnosis of OAB or urinary retention. The model coefficient estimate for each statistically significant fixed effect is given in Table 2. The equation used to generate the estimates is given in Supplemental Equation 1. Patient demographic data (BMI, age, or sex) did not have a significant fixed effect. The relationship between EAS response and applied current across pulse widths and diagnoses can be seen in Figure 2. The EAS AUC for each observed trial is shown in Supplemental Figure 4, segmented by each diagnosed condition, highlighting the overlap between OAB and retention diagnoses. Overall, we included 343 monopolar trials across sixteen participants in this model.

**Table 2.** Linear mixed model estimating anal sphincter activation. Summary of the significant fixed effects for the linear mixed model estimating EAS AUC(µV*s). The data includes all monopolar trials (N = 343 trials from 16 participants). A representative equation for the corresponding linear mixed model is shown in Supplemental Equation S1.

| Fixed Effect | Estimate | Standard Error | 95% Confidence Interval | p value |
| --- | --- | --- | --- | --- |
| Intercept | 2.73 | 0.69 | (1.38, 4.08) | 4.67e-4 |
| Current | 0.79 | 0.10 | (0.60, 0.98) | 1.5e-14 |
| Pulse Width 210 $\mu\text{s}$ | 1.91 | 0.35 | (1.23, 2.61) | 8.27e-8 |
| Pulse Width 450 $\mu\text{s}$ | 2.38 | 0.39 | (1.60, 3.15) | 4.22e-9 |
| OAB yes | -2.79 | 0.58 | (-3.91, -1.66) | 2.66e-4 |
| Retention yes | -2.28 | 0.56 | (-3.39, -1.17) | 8.94e-4 |

OAB: overactive bladder EAS: external anal sphincter AUC: area under the curve

**Figure 2.**
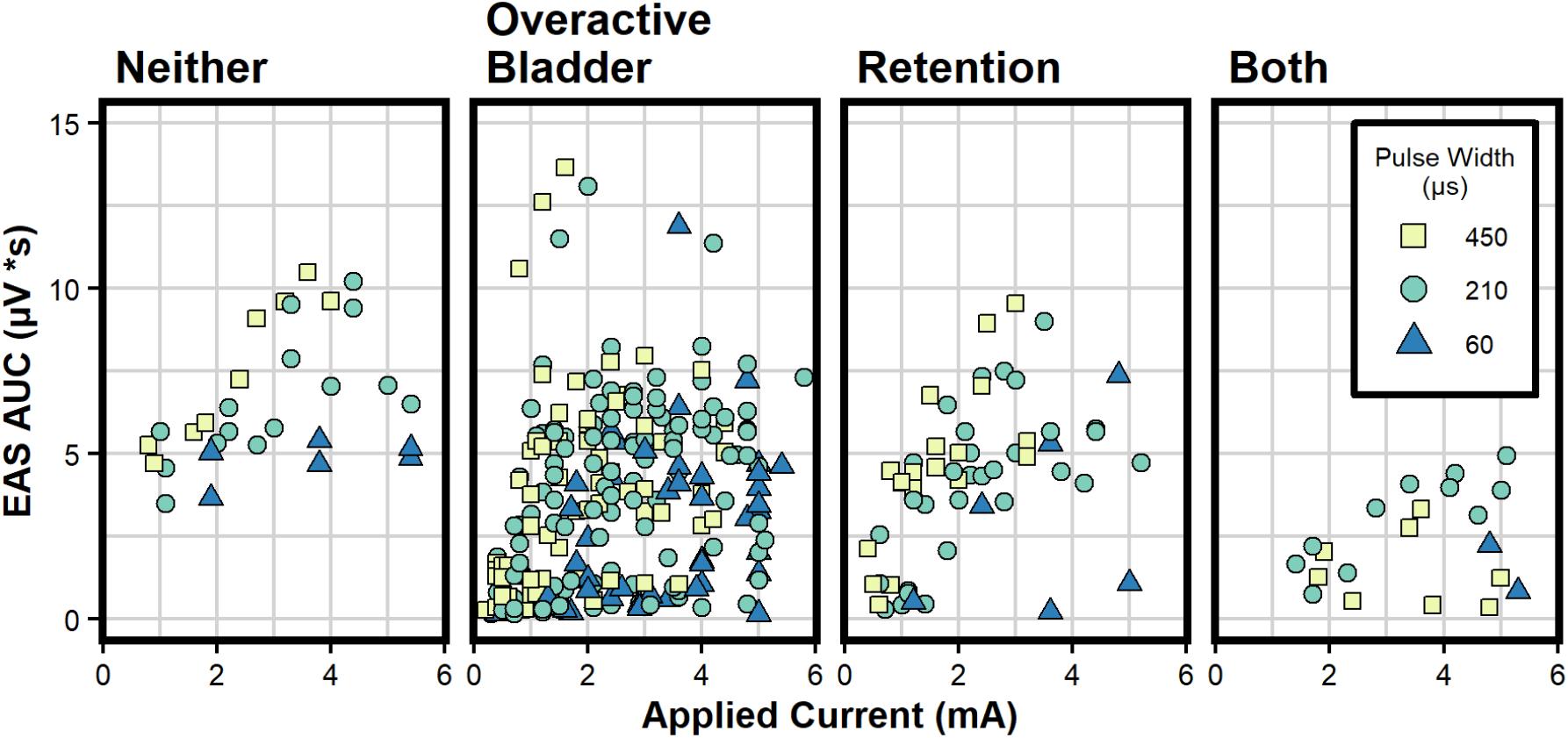
Anal sphincter response magnitude plotted against applied current for all monopolar trials across all participants. Data is separated by participant diagnosis of overactive bladder and/or urinary retention. Each trial was performed with a pulse width of 60, 210, or 450 µs and is labeled according to the legend.

Eleven study participants (all female) had fluoroscopy and CT images that allowed for identifying lead migration distances. Five participants (all female) had a lead migration distance less than 5 mm, and their corresponding electrode-to-nerve distances are shown in Figure 3. We created two linear mixed models to investigate the impact of stimulation parameters on EAS AUC and EAS threshold for participants with valid electrode-to-nerve distances. The estimates for the fixed effect of applied current, pulse width, and electrode-to-nerve distance on EAS AUC were all statistically significant and are shown in Supplemental Table 1. Additionally, when controlling for other variables, each millimeter between the nerve and electrode decreased the expected EAS AUC by 0.27 µV*s (95% confidence interval [CI] = -0.41 to -0.13; p = 0.0011). A second mixed model showed that electrode-to-nerve distance and pulse width both significantly affect EAS threshold (Supplemental Table 2). Each millimeter between the nerve and electrode increased the expected threshold by 0.22 mA (95% CI = 0.15 to 0.29; p = 2.7e-7). We performed linear regressions between the average electrode-to-nerve distance and change in each survey score. There was a significant relationship (p = 0.049) between AUASI score change and average electrode-to-nerve distance, indicating greater bladder symptom improvement with reduced electrode-to-nerve distance, but not with any other survey score change (Figure 4).

**Figure 3.**
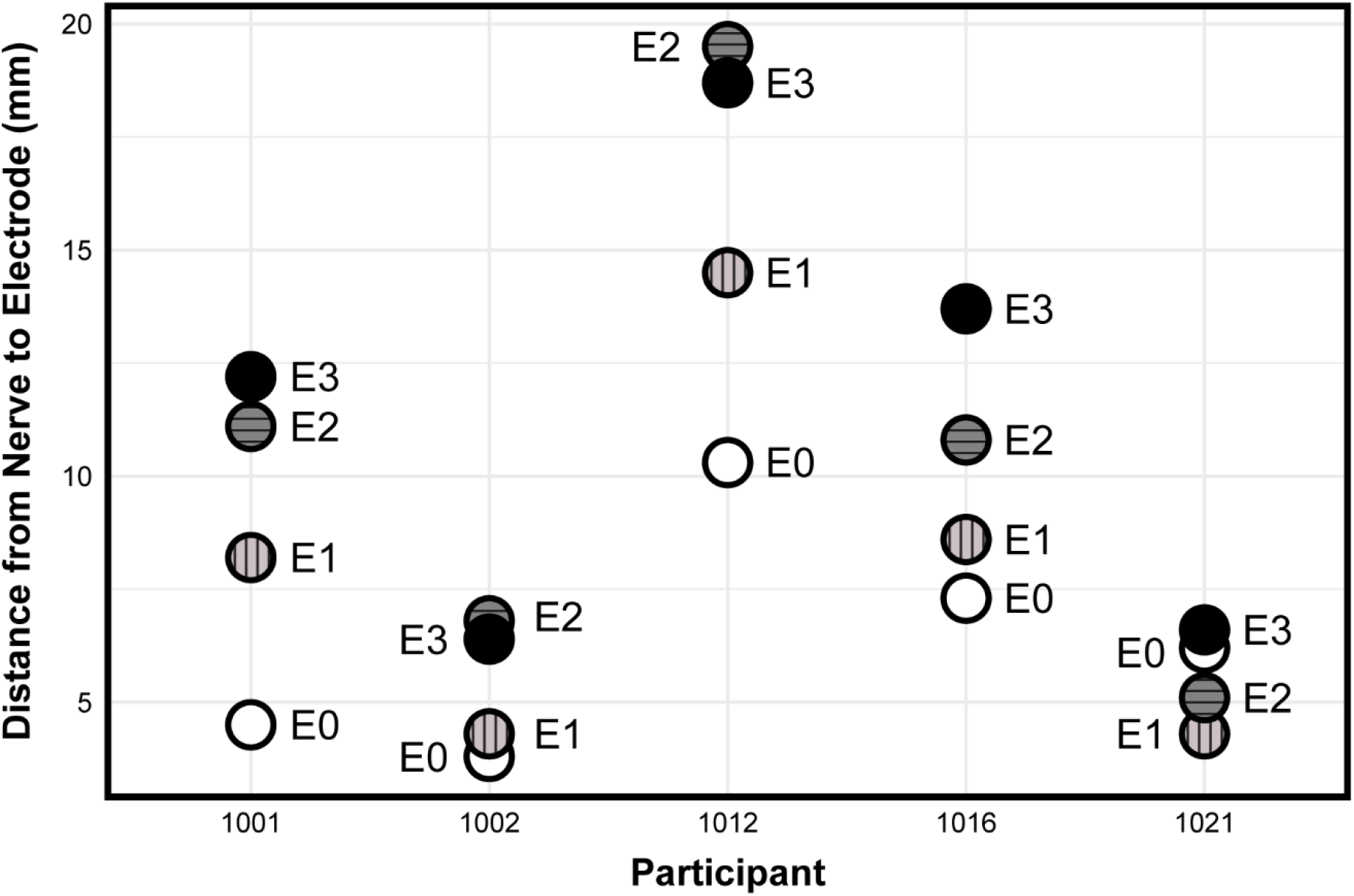
Measured distances (in millimeters) between each electrode on the implanted quadripolar lead and the closest point of the pudendal nerve for the five participants with minimal lead migration (<5 mm). Electrodes are labeled from E0 to E3 with E0 representing the electrode most distal to the implantable pulse generator to E3 representing the electrode most proximal to the implanted lead.

**Figure 4.**
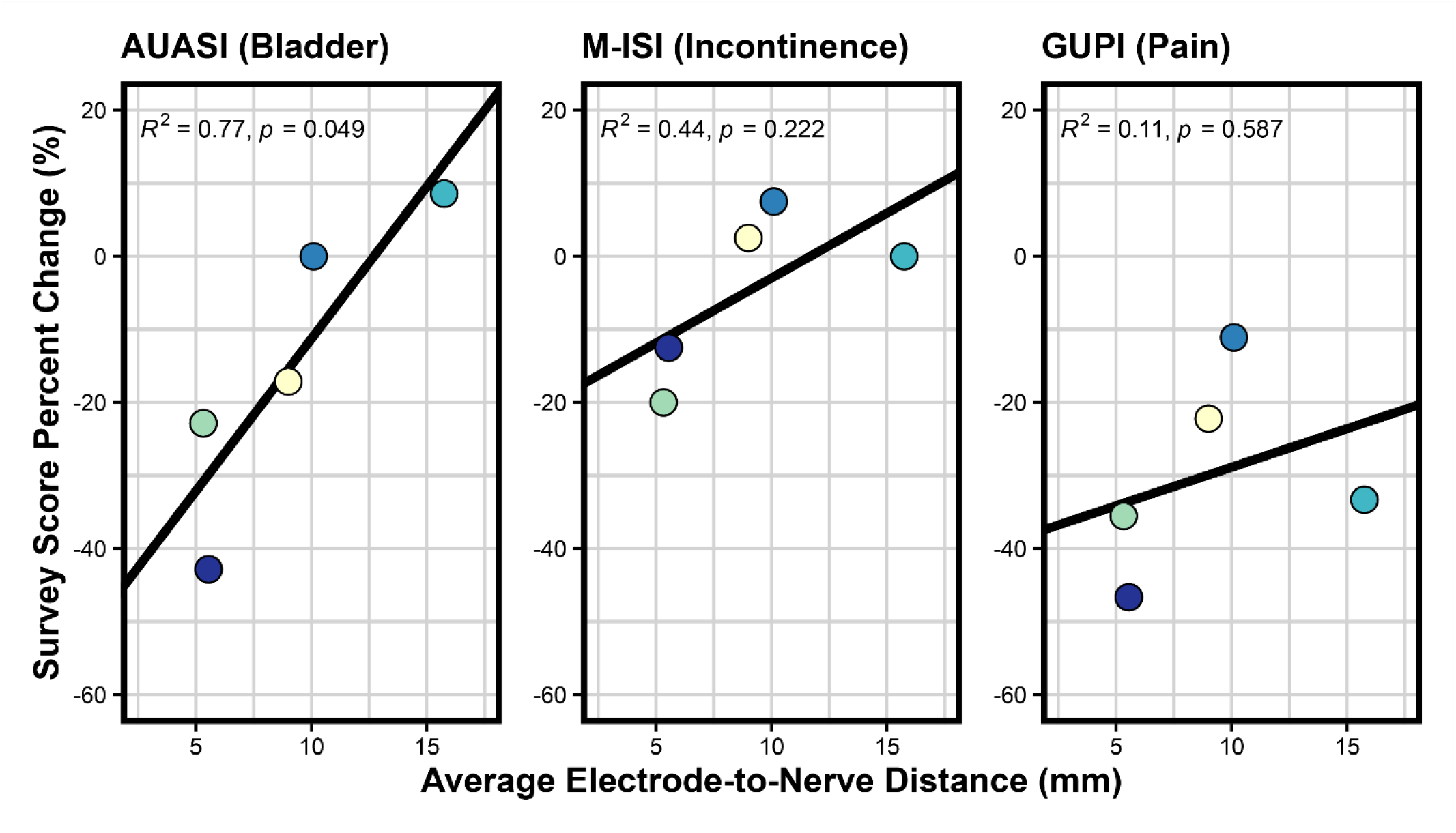
Linear regression fits between each participant’s average electrode-to-nerve distance and symptomatic survey score percent change before and after receiving pudendal neuromodulation treatment. Surveys on asymptomatic functions (CRAD-8 (Bowel), FSFI (Female sexual function)) did not have relationships to electrode-to-nerve distance (p = 0.606, 0.706, respectively).

## Discussion

In this observational study, we examined the influence of key patient data and stimulation parameters on pudendal nerve recruitment. To our knowledge, this is the first study which investigates these relationships in human subjects.

A pulse width of 210 µs is the current standard of care for pudendal neuromodulation[27] but there is little documentation supporting this choice, particularly in human subjects. We show that when compared to 60 µs and 450 µs, a pulse width of 210 µs balances the minimum required current (Figure 1A) and charge (Figure 1B) and therefore requires less energy to recruit the pudendal nerve. These results agree with the findings of an earlier study [28] that showed a pulse width of 200 µs minimizes the energy required to activate afferent fibers of the pudendal nerve and align with the most commonly used sacral neuromodulation pulse width of 210 µs [29] supporting the continued use of common sacral neuromodulation stimulation pulse widths in pudendal neuromodulation. Thus, our findings confirm that pulse width does not need to be adjusted for pudendal nerve stimulation, and clinicians can focus on amplitude and frequency when determining stimulation parameters.

We estimated the relationship between EAS activation and age, BMI, and LUTD diagnosis with a linear mixed model. Age, sex, BMI and diagnosis of incontinence or pelvic pain did not significantly affect anal sphincter recruitment.

However, diagnosis of OAB or retention significantly reduced the EAS activation based on 95% confidence interval testing (Table 2). Participants with diagnoses of OAB or retention had lower EAS activation across all applied currents (Figure 2). If true, these results would suggest that people with these diagnoses might require more current than is typical to activate the EAS and recruit the pudendal nerve. However, it is important to note that there was an unbalanced distribution of diagnoses in our study population and diagnoses of neither OAB nor retention (n=2) or both OAB and retention (n=1) were rare. This limited the amount of data collected for these cases, shown in Supplemental Figure 4, and implies that the effects of certain diagnoses were mainly dependent on a few participants, limiting the generalizability of our results. Also, the model estimates for OAB and retention had large errors of 0.58 and 0.57, respectively, showing that the strength of the effect varied across participants. The model also showed that pulse width and current significantly impact EAS recruitment (Table 2). We observed that larger electrode-to-nerve distances were significantly related to reductions in the magnitude of the EAS response and increases in the current required to activate the EAS at threshold, as is shown in Supplemental Tables 1 and 2 and aligning with bioelectric principles. Therefore, examining changes in the EAS EMG AUC and EAS activation thresholds during intraoperative stimulation could inform estimates of the relative electrode-to-nerve distance and help clinicians optimize lead placement.

We observed a significant relationship between electrode-to-nerve distance and AUASI scores (Figure 4), with improved bladder outcomes when the distance was minimized. We also observed that six of eleven participants had lead migration larger than 5 mm between the stage-1 implant surgery and CT imaging. Examination of lead migration was not a primary goal of this study, however it is not commonly reported in the literature. Future studies should specifically investigate whether lead migration has a significant relationship to outcomes or changes in pelvic floor recruitment.

Generally, the goal when implanting a pudendal stimulation lead is to activate the anal sphincter with a current below 2 mA [30] with at least three electrodes [31]. In this study, 94% of participants had EAS activation below 2 mA with a 210 µs pulse width (Supplemental Figure 2). While lead implantation at the pudendal nerve is a challenging procedure, these results show that pudendal nerve stimulation can consistently activate the EAS at low current when the lead is placed by an experienced surgeon. During the implant surgery, monopolar stimulation is used to locate the nerve and adjust the position of the foramen needle or lead. However, most pudendal neuromodulation patients use bipolar electrode combinations to manage their symptoms at home. In our data, we did not see a significant difference in the threshold current for monopolar trials and bipolar trials, shown in Supplemental Figure 3, supporting the continued use of monopolar stimulation to confirm intraoperative lead location. We provide evidence supporting the typical use of electrodes with the lowest EAS activation threshold when performing patient programming, as our results show these electrodes are likely closest to the nerve and therefore are expected to deliver the most effective therapy.

It’s important to note some limitations in this study. We did not perform an a priori power analysis to support our target sample size due to a lack of relevant effect sizes and fixed capabilities in patient recruitment. However, post hoc power analyses suggest that we had sufficient ability to detect our observed findings. We reduced the estimate of each linear mixed model fixed effect by 20%, following good principles to avoid computing the observed power[32] and taking a more conservative approach than a 15% reduction used previously [33], and obtained a power of over 90% for each variable in the AUC magnitude model with 16 participants, using the simr package in R[34]. Similarly, for the linear mixed model that related electrode-to-nerve distance to AUC magnitude using data from 5 participants, we obtained a power of 84.8%. Evoked EAS responses due to stimulation were monitored live and identified manually, leaving room for human judgement errors and misidentification of threshold current. However, as the amplitude step interval was only 0.1 mA, we expect any threshold errors were minimal. The relative depth of the EAS needles was not tracked across procedures. In general, the surgeon seeks to place the needles as deep as possible with a consistent position on opposite sides of the sphincter, however it is possible one or both needles shifted during a surgical procedure which could affect the EMG recording. Along with a relatively small sample size, there was an unbalanced grouping in the number of participants with OAB and the gender ratio of participants with OAB, which limit the generalizability of our conclusions. Additional studies should be done with larger, balanced samples sizes and diagnoses to investigate the relationship between EAS recruitment and OAB diagnosis.

This study emphasizes the benefits of a 210 µs stimulation pulse width, as compared to 60 and 450 µs, and provides support for its continued use in pudendal neuromodulation. Pudendal nerve stimulation is effective at activating the pudendal nerve and EAS in patients with a wide range of LUT symptoms and backgrounds. Minimizing the distance between the nerve and electrode is critical for recruitment of the pudendal nerve. Our results show that readily available physiological measurements like EAS threshold and AUC magnitude may be good indicators of electrode-to-nerve distance and useful for determining when that distance is minimized. Larger distances require more current to activate the nerve and may therefore reduce the battery lifespan of implanted devices and reduce the size of the therapeutic window. Future studies can utilize the data collected in this work towards a priori sample size calculations and more comprehensive designs. Additionally, future studies can more accurately determine the relationship between electrode-to-nerve distance and observed AUC response, examine how bipolar stimulation affects pudendal nerve recruitment at different distances from the electrode to the nerve, and look deeper at how LUT dysfunction may affect recruitment of the EAS with pudendal nerve stimulation. Finally, this study supports that pudendal neuromodulation has a role in treating refractory overactive bladder and pelvic pain. The relationships observed here may be similar in sacral neuromodulation, and analogous studies with patients receiving SNM are warranted.

## Data Availability

The data set from the study will be found on the SPARC Science data portal at DOI: 10.26275.pc8r-r3iu after completing the SPARC curation process.

https://doi.org/10.26275/pc8r-r3iu

## Abbreviations

EAS: External Anal Sphincter
LUT: Lower Urinary Tract
LUTD: Lower urinary tract disorders
CT: Computed tomography
MRI: Magnetic resonance imaging
AUASI: American Urological Association Symptom Index
M-ISI: Michigan Incontinence Symptom Index
FSFI-6: Female Sexual Function Index-6
SHIM: Sexual Health Inventory for Men
SHIM CRAD-8: Colorectal-Anal Distress Inventory-8
FGUPI: Female Genitourinary Pain Index
MGUPI: Male Genitourinary Pain Index
EMG: Electromyography
AUC: Area under the curve
IQR: Interquartile range
OAB: overactive bladder

## Acknowledgments

We thank Vanessa Pruitt and Mackenzie Moore for their contributions as study coordinators and Gaurang Shah for his help in reviewing MRI scans. We also thank Kelly Ridenour and Stephanie Jones for their EMG identification expertise in the operating room. This project was funded by an award from the National Institutes of Health SPARC program (OT2OD028191).

