## Supplemental Information for "Factors affecting anal sphincter recruitment during intraoperative pudendal nerve stimulation: an observational study"

### Title


### Supplemental Information

Supplemental Table 1: The significant fixed effects on EAS EMG AUC (µV*s) in the linear mixed model incorporating electrode-to-nerve distance. The data includes all monopolar trials with electrode-to-nerve distance data (N=103 trials from 5 participants).

| Fixed Effect | Estimate | Standard Error | 95% Confidence Interval | p-value |
|---|---|---|---|---|
| Intercept | 0.76 | 0.73 | (-0.68, 2.19) | 0.32 |
| Current | 0.90 | 0.15 | (0.60, 1.20) | 7.3e-8 |
| Pulse Width 210 µs | 2.69 | 0.50 | (1.71, 3.67) | 5.6e-7 |
| Pulse Width 450 µs | 2.60 | 0.57 | (1.44, 3.67) | 2.2e-5 |
| Distance | -0.27 | 0.07 | (-0.41, -0.13) | 1.1e-3 |

Supplemental Table 2: The significant fixed effects on EAS threshold (mA) in the linear mixed model incorporating electrode-to-nerve distance data. The data includes all monopolar trials with electrode-to-nerve distance data (N=35 trials from 5 participants).

| Fixed Effect | Estimate | Standard Error | 95% Confidence Interval | p-value |
|---|---|---|---|---|
| Intercept | 0.78 | 0.35 | (0.11, 1.46) | 0.030 |
| Pulse Width 210 µs | -0.86 | 0.32 | (-1.49, -0.23) | 0.011 |
| Pulse Width 450 µs | -1.47 | 0.36 | (-2.17, -0.77) | 2.8e-4 |
| Distance | 0.22 | 0.03 | (0.15, 0.29) | 2.7e-7 |

Supplemental Equation 1: The equation relating the statistically significant factors of interest to the observed EAS response magnitude, written in R lme4 format, to generate the estimates shown in Table 2.

$$\boldsymbol{EAS\ AUC \sim Pulse\ Width + Current + OAB + Retention + (1|Participant)}$$

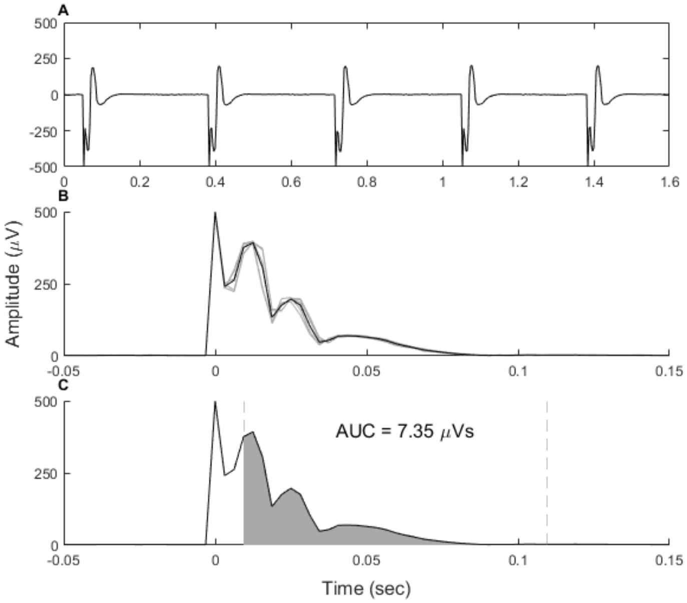


Supplemental Figure 1: Anal sphincter EMG and AUC Quantification procedure. (A) Example EAS EMG after signal filtering showing five consecutive responses during a single trial. Stimulation was applied at 3 Hz. (B) Each individual response in A is segmented, rectified, and aligned at the stimulation artifact. Individual traces in are in gray and the averaged signal is in black. (C) The shaded area under the rectified average EMG trace, between the vertical dashed lines, is the calculated area under the curve (AUC).

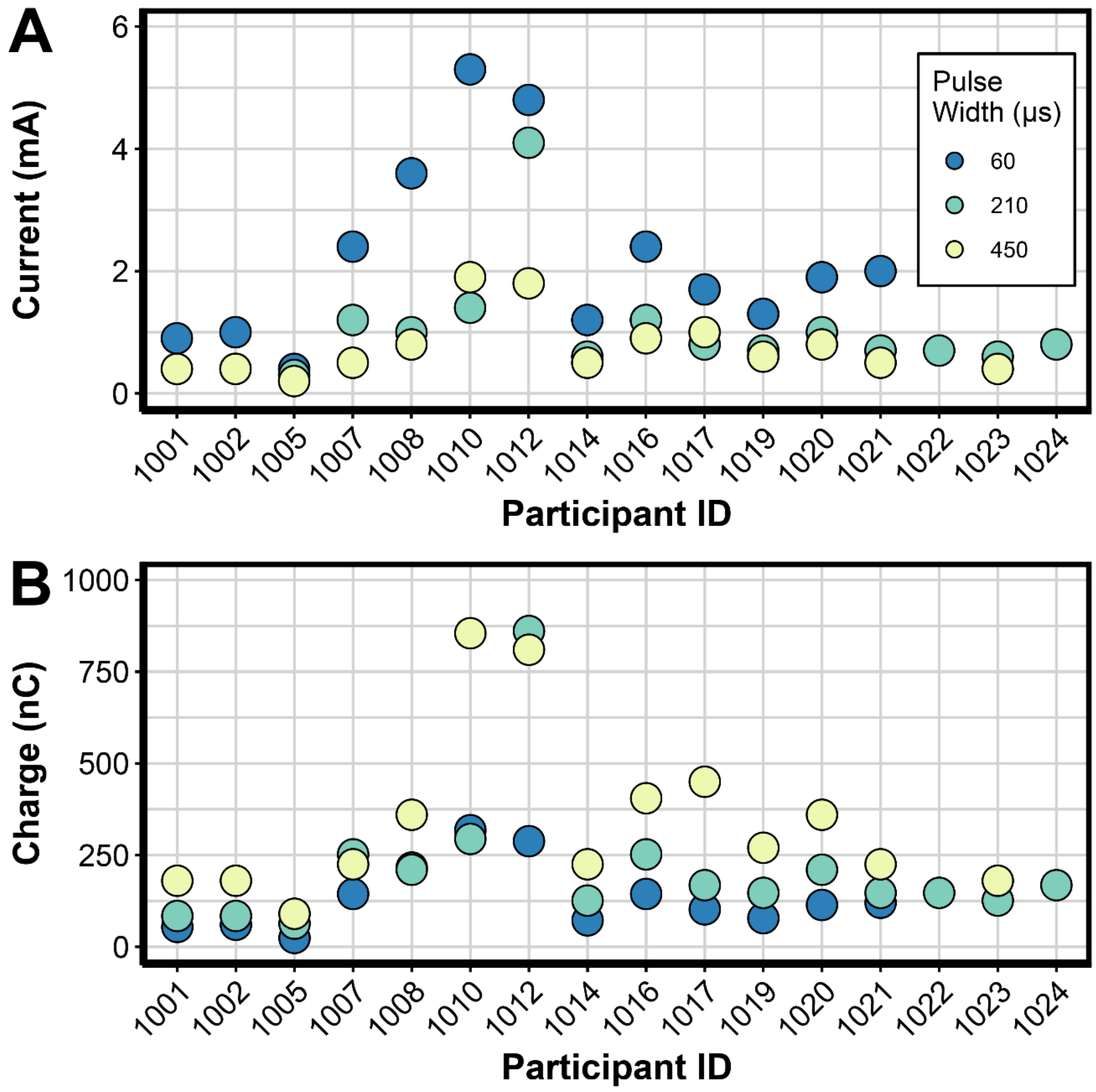


Supplemental Figure 2: Monopolar threshold current and charge for all participants at each tested pulse width. A) EAS threshold current for each participant. If we tested the threshold for multiple electrodes at a given pulse width then the lowest threshold is shown. B) The charge at EAS threshold for each participant.

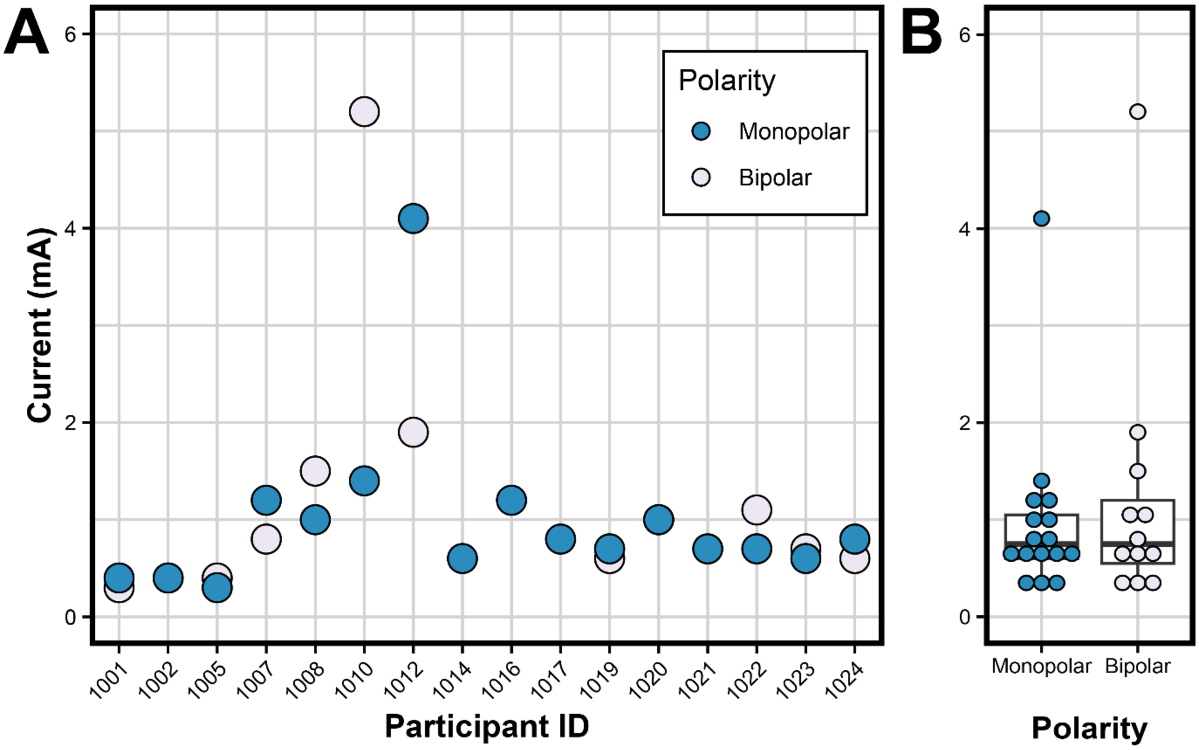


Supplemental Figure 3: EAS activation thresholds for 210 µs pulse width monopolar and bipolar trials. A) The minimum EAS threshold for monopolar and bipolar stimulation is shown for each participant. B) Aggregated EAS thresholds across all participants for monopolar and bipolar trials. Box plots represent the inter-quartile range (25%-75%; IQR) for the height of each box, with the bold central line indicating the median. Tips of whiskers extend to the value(s) within 1.5 x IQR of each box side.

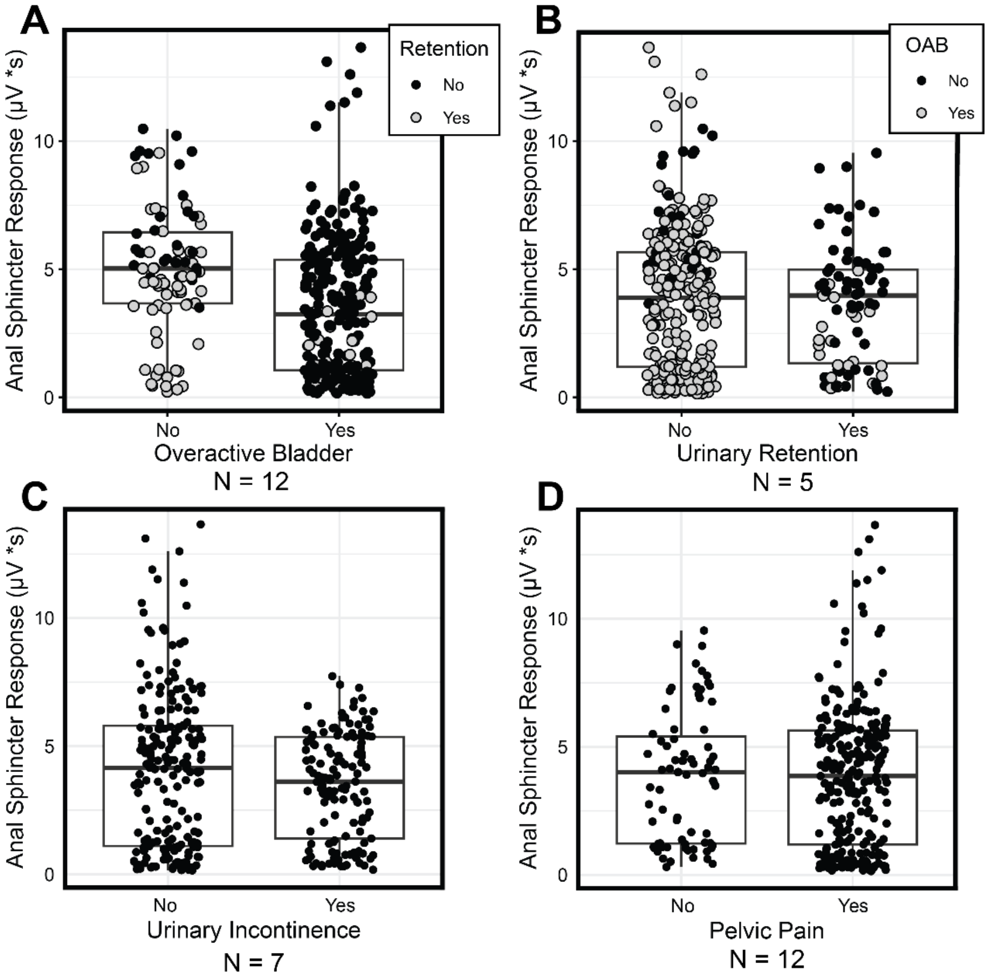


Supplemental Figure 4: The EAS response magnitude dependence on participant diagnosis of A) Overactive Bladder, B) Urinary Retention, C) Urinary Incontinence, or D) Pelvic Pain. The N value under each x-axis indicates the number of participants diagnosed with each condition out of 16 total participants. The legends in sub-figures A and B show comorbidities of Retention or Overactive Bladder, respectively.
